# A Bayesian computational model reveals a failure to adapt interoceptive precision estimates across depression, anxiety, eating, and substance use disorders

**DOI:** 10.1101/2020.06.03.20121343

**Authors:** Ryan Smith, Rayus Kuplicki, Justin Feinstein, Katherine L. Forthman, Jennifer L. Stewart, Martin P. Paulus, Tulsa 1000 investigators, Sahib S. Khalsa

## Abstract

Recent neurocomputational theories have hypothesized that abnormalities in prior beliefs and/or the precision-weighting of afferent interoceptive signals may facilitate the transdiagnostic emergence of psychopathology. Specifically, it has been suggested that, in certain psychiatric disorders, interoceptive processing mechanisms either over-weight prior beliefs or under-weight signals from the viscera (or both), leading to a failure to accurately update beliefs about the body. However, this has not been directly tested empirically. To evaluate the potential roles of prior beliefs and interoceptive precision in this context, we fitted behavior in a transdiagnostic patient population on an interoceptive awareness (heartbeat tapping) task to a Bayesian computational model. Modeling revealed that, during an interoceptive perturbation condition (inspiratory breath-holding during heartbeat tapping), healthy individuals (N=52) assigned greater precision to ascending cardiac signals than individuals with symptoms of anxiety (N=15), depression (N=69), co-morbid depression/anxiety (N=153), substance use disorders (N=131), and eating disorders (N=14) – who failed to increase their precision estimates from resting levels. In contrast, we did not find strong evidence for differences in prior beliefs. These results provide the first empirical computational modeling evidence of a selective dysfunction in adaptive interoceptive processing in psychiatric conditions, and lay the groundwork for future studies examining how reduced interoceptive precision influences body regulation and interoceptively-guided decision-making.

**Author Summary:** Interoception is the process by which the nervous system senses the internal state of the body. It provides the brain with important information to adaptively guide the regulation of both internal body states and behavior. Interoceptive dysfunction is thought to play a role in multiple psychiatric disorders. Theoretical models propose that the computational mechanisms of interoceptive dysfunction are caused by overly precise prior beliefs about body states (“hyperprecise priors”) or underestimates of the reliability of the information carried by ascending signals from the body (“low sensory precision”). Our empirical approach tested for evidence of these mechanisms across several psychiatric disorders, using a computational model of perception during performance of a heartbeat perception task. We found evidence of low sensory precision within individuals with anxiety, depression, eating disorders, and/or substance use disorders, relative to healthy individuals. This difference occurred only during a breath-holding condition designed to enhance heartbeat signals. We did not find evidence for hyperprecise priors in the patient groups. The data from this study support the argument for computational mechanisms of interoceptive dysfunction across several psychiatric disorders, and suggests that these conditions may be characterized by an inability to adjust sensory precision when signals from the body change.

## Introduction

Interoception plays an important role in a number of psychiatric disorders. Interoceptive dysfunction has been observed in depression, anxiety, eating, and substance use disorders, among others (reviewed in (1)). For example, depressed patients exhibit reduced accuracy when asked to count their own heartbeats (2-4), and counting accuracy is negatively correlated with depressive symptoms (5). Several studies have also shown heightened interoceptive sensations in panic disorder under high arousal states (reviewed in (6)). Other studies have reported evidence of blunted neural responses during interoceptive processing in substance users (7), and that patients with eating disorders show stronger expectation effects on interoception during modulations of arousal (8). However, the mechanisms underlying these potential interoceptive dysfunctions in psychiatric disorders have not yet been demonstrated in empirical work.

In recent years, a growing body of theoretical work within neuroscience and psychiatry has begun to highlight plausible neurocomputational accounts of interoceptive processing (9-15), as well as accounts of exteroceptive (16, 17), cognitive (18-26), emotional (27, 28), and motor control (29-31) functions that plausibly interact with interoception. In some cases, empirical studies have also found support for computational models and computational abnormalities in relation to specific psychiatric symptoms/disorders (26, 32-34). With respect to sensory processing, which has focused to a large degree on neural models of approximate probabilistic (Bayesian) inference, empirical results in this area have come almost exclusively from the exteroceptive domain. And while recent work has begun to test qualitative predictions of Bayesian models of interoception (35), no study to our knowledge has yet explicitly fit computational models to brain or behavioral responses during experimental modulations of interoception in individuals with psychiatric disorders.

Drawing on this theoretical literature, we and others (9, 13-15, 36-38) have previously proposed that symptoms of multiple psychiatric conditions could be explained by faulty interoceptive computational processes (and associated visceromotor control processes), due to an inappropriate weighting of prior beliefs and sensory evidence. Although these models of computational dysfunction in interoception could have broad explanatory power, the putative mechanisms they propose have yet to be empirically tested using formal computational models. In this paper, we use a formal Bayesian computational model of perception to examine whether there is evidence for the transdiagnostic dysfunction of interoceptive processing suggested in these previous proposals within the cardiovascular system.

Historically, the most common empirical paradigms for studying interoception have focused on the perception of cardiac signals. Common heartbeat perception tasks include those involving heartbeat counting (39), heartbeat tapping (40), and other means of assessing cardiac interoception (41-45). While each approach has certain limitations (46-50), one consistent finding is that cardiac interoceptive awareness is quite poor in the majority of participants tested during resting conditions – where only roughly 35% of individuals appear to accurately perceive their own heartbeats (6). In contrast, when visceral states are perturbed, cardiac perception becomes more accurate. This is particularly true for interoceptive accuracy under conditions of heightened cardiorespiratory arousal (51-53).

Active inference models, and Bayesian predictive processing models more generally, have been the main computational framework within which interoceptive dysfunction has been discussed (13, 38, 54-56). Several authors have suggested that one important transdiagnostic factor within mental disorders may be an inability of the brain to update its model of the body in the face of interoceptive prediction errors (i.e., mismatches between expected and received afferent interoceptive signals from the body). In predictive coding and active inference models, this kind of aberrant belief updating is thought to come about through a dysfunctional “precision weighting” mechanism, which governs the relative influence of prior beliefs and afferent bodily signals in determining perception (and informing visceral regulation). Simply put, it is suggested that, across multiple mental health conditions, the brain may treat afferent bodily signals (and associated prediction errors) as though they are not reliable indicators of bodily states during interoceptive inference – leading perception to be insufficiently constrained by true visceral states and primarily determined by (in many cases maladaptive) prior beliefs. Misestimating the state of the body could in turn promote a number of transdiagnostic symptoms. For example, interoceptive feelings are intimately tied to emotions (57-59); poor body perception (e.g., high uncertainty about internal physiological conditions and a resulting inability to efficiently regulate them) could thus maintain unpleasant emotional states. Chronic underestimates of available metabolic resources may contribute to apathy and anhedonia (56), and overestimates of the evidence uncomfortable bodily sensations provide for physical threat (e.g., a heart attack) may contribute to anxiety and panic (60, 61).

It’s important to emphasize, however, that computational models often include several additional parameters. Aside from the precision-weighting of sensory signals, individuals can also have differences in (for example) prior beliefs about what they will perceive (assumed in any Bayesian model of perception; e.g., predictive coding (62)), and differences in how quickly they update those prior beliefs over repeated observations (i.e., “learning rate”; i.e., which can be dependent upon the relative precisions of sensory signals and prior beliefs). Formal computational models are often necessary to distinguish which parameters show differences between individuals and best explain differences in perception. Thus, there are multiple computational mechanisms that could account for individual differences in interoception in clinical populations. One major goal in computational psychiatry is to “computationally phenotype” patients by identifying which sets of a parameter values best account for their neural and behavioral responses (including self-reported perceptual experience) and use this information to guide treatment (20, 22, 25).

In the present study, we apply a novel computational phenotyping approach, using a Bayesian model of perception, to identify the computational parameters that best explain behavior on a cardiac perception (heartbeat tapping) task performed by a transdiagnostic clinical sample of individuals with psychiatric disorders as well as a healthy comparison (HC) sample. Given previous findings that performance on cardiac perception tasks can be significantly altered by different task instructions ((63); i.e., potentially by adjusting prior beliefs), we chose to assess task performance under two resting conditions in which participants were instructed to apply different confidence thresholds: 1) a condition where guessing was allowed and 2) a condition in which they should not guess (i.e., they should only tap a key if they were sure they actually felt a heartbeat). Given the notably poorer cardiac perception of most human beings at rest compared to during altered arousal states (6), in addition to the resting conditions we also chose to assess individual differences in cardiac perception during a non-invasive interoceptive perturbation (a breath-hold) condition (which also included the no-guessing instruction) that was expected to increase the precision of the afferent cardiac signal and improve cardiac perception above floor values in a greater number of individuals (i.e., we expected that cardiac perception would be generally poor during resting conditions, and that the breath-hold condition would result in improved performance on average). Thus, comparing the guessing and no-guessing conditions allowed assessment of the effects of altering prior beliefs with different task instructions, and comparison of the no-guessing a breath-hold conditions allowed assessment of changes in afferent signal precision under identical (i.e., no-guessing) task instructions. Our primary aims were to 1) demonstrate the sensitivity of our novel computational approach in measuring the condition-specific precision-weighting of interoceptive signals and prior beliefs across a transdiagnostic sample of individuals with depression, anxiety, substance use disorders, and/or eating disorders, 2) test the hypothesis (as previously proposed; e.g., (10, 38, 56)) that these patient groups would show lower interoceptive precision weightings than HCs, more precise prior beliefs than HCs, or both, and 3) establish whether prior beliefs and/or interoceptive precision is abnormal in general (relative to healthy participants) or selectively within resting or interoceptive perturbation conditions.

## Methods

### Participants

Data were collected from 500 participants (153 male) as a part of the Tulsa 1000 (T1000) project, a naturalistic longitudinal study that recruited subjects based on the dimensional NIMH Research Domain Criteria framework (a full description of this pre-planned project can be found in (64)). Individuals aged 18-55 years were screened on the basis of dimensional psychopathology scores. Inclusion was based on the following measures: Patient Health Questionnaire (PHQ-9; (65)) ≥ 10, Overall Anxiety Severity and Impairment Scale (OASIS; (66)) ≥ 8, Drug Abuse Screening Test (DAST-10; (67)) score > 2, and/or Eating Disorder Screen (SCOFF; (68)) score ≥ 2 (for screening measure scores, see **Table 1**). HCs who did not show elevated symptoms or psychiatric diagnoses were also included. Participants were excluded if they (i) tested positive for drugs of abuse, (ii) met criteria for psychotic, bipolar, or obsessive-compulsive disorders, or reported (iii) history of moderate-to-severe traumatic brain injury, neurological disorders, or severe or unstable medical conditions, (iv) active suicidal intent or plan, or (v) change in psychotropic medication status within 6 weeks. Full inclusion/exclusion criteria are described in (64). The study was approved by the Western Institutional Review Board. All participants provided written informed consent prior to completion of the study protocol, in accordance with the Declaration of Helsinki, and were compensated for participation. ClinicalTrials.gov identifier: #NCT02450240.

**Table 1.** Mean (and standard deviation) for clinical and demographic variables.

| Individual difference variable* | Healthy comparison (N = 52) | Anxiety (N = 15) | Depression (N = 69) | Depression + Anxiety (N = 153) | Eating Disorder (N = 14) | Substance use disorder (N = 131) | p-value |
| --- | --- | --- | --- | --- | --- | --- | --- |
| Age | 32.04 (11.08) | 36.42 (10.01) | 37.12 (11.83) | 35.28 (11.23) | 27.40 (9.83) | 34.03 (8.88) | 0.012 |
| Sex | 50% male | 33% male | 30% male | 26% male | 14% male | 45% male | 0.001 |
| PHQ-9 | 0.83 (1.29) | 7.47 (5.80) | 13.48 (4.73) | 13.10 (5.21) | 12.93 (7.98) | 6.68 (5.91) | <0.001 |
| OASIS | 1.37 (1.89) | 10.60 (2.16) | 7.55 (3.34) | 10.51 (3.12) | 9.50 (4.93) | 5.94 (4.77) | <0.001 |
| DAST-10 | 0.10 (0.30) | 0.33 (0.49) | 0.72 (1.66) | 0.58 (1.13) | 1.23 (2.62) | 7.55 (2.05) | <0.001 |
| PTT | 0.20 (0.02) | 0.20 (0.01) | 0.19 (0.01) | 0.20 (0.02) | 0.20 (0.02) | 0.20 (0.02) | 0.261 |
| **BMI** | 27.59<br>(5.54) | 27.05<br>(6.02) | 28.51<br>(5.47) | 28.77<br>(5.49) | 22.25<br>(4.43) | 28.23<br>(4.56) | 0.001 |

\*PHQ-9 = Patient Health Questionnaire 9; OASIS = Overall Anxiety Sensitivity and Impairment Scale; DAST-10 = Drug Abuse Screening Test; PTT = median pulse transit time; BMI = Body Mass Index.

Similar to previous studies of the T1000 cohort (69), participants were grouped based on DSM-IV or DSM-5 diagnosis using the Mini International Neuropsychiatric Inventory 6 or 7 (MINI; (70)), and included those with major depressive disorder only (DEP), an anxiety disorder only (ANX; social anxiety, generalized anxiety, panic, or posttraumatic stress disorder), comorbid MDD and anxiety disorder (DEP+ANX), substance use disorders (SUDs; recreational drugs excluding alcohol and nicotine; with or without comorbid anxiety and mood disorders), eating disorders (EDs; with or without comorbid anxiety and mood disorders), and HCs with no mental health diagnoses. In this study we examined all groups from the first 500 participants of the T1000 project (recruited from January 5, 2015 to February 22, 2017).

### Heartbeat perception task

As part of the T1000 project, participants completed a large number of assessments, self-report measures, and behavioral tasks (detailed in (64)). Here we focus on data from a cardiac perception task on which we have previously reported (i.e., on a subset of the participants reported here, with analyses unrelated to computational modeling (71, 72)), wherein participants were asked to behaviorally indicate the times at which they felt their heartbeat. The utilization of the heartbeat tapping measure as an index of perception was based on a previously developed heartbeat tapping task (40); for a more recent example, see (73)). The task was repeated under multiple conditions designed to assess the influence of cognitive strategy and physiological perturbation on performance. In the initial task condition, participants were simply instructed to close their eyes and press down on a key when they felt their heartbeat, to try to mirror their heartbeat as closely as possible, and even if they weren’t sure they should take their best guess (the “guessing” condition). Participants completed this (and each other) task condition over a period of 60 seconds. In the second task condition, all instructions were identical except that they were told to only press the key when they actually feel their heartbeat, and if they do not feel their heartbeat then they should not press the key (the “no-guessing” condition). In other words, unlike the first time they completed the task, they were specifically instructed not to guess if they didn’t feel anything, which could be understood as altering prior beliefs associated with confidence thresholds (i.e., higher confidence was required to choose to tap than in the guessing condition, where this change in prior confidence thresholds could differ between individuals).

Finally, in the perturbation condition, participants were again instructed not to guess but were also asked to first empty their lungs of all air and then inhale as deeply as possible and hold it for as long as they could tolerate (up to the length of the one-minute trial) while reporting their perceived heartbeat sensations. This third condition (the “breath-hold” condition) was used in an attempt to putatively increase the strength of the afferent cardiac signal by increasing physiological arousal. We expected that cardiac perception would be poor in the guessing condition, that tapping would be more conservative in the no-guessing condition, and that the breath-hold condition would result in improved performance on average. As a control condition, we also included an identical task where participants were instructed to tap every time they heard a 1000Hz auditory tone presented for 100ms (78 tones, randomly jittered by +/- 10% and presented in a pattern following a sine curve with a frequency of 13 cycles/minute, mimicking the range of respiratory sinus arrhythmia during a normal breathing range of 13 breaths per minute). This was completed between the first (guessing) and second (no-guessing) heartbeat tapping conditions.

Directly after completing each task condition, individuals were asked the following using a visual analogue scale:

“How accurate was your performance?”
“How difficult was the previous task?”
“How intensely did you feel your heartbeat?”
Each scale had anchors of “not at all” and “extremely” on the two ends. Numerical scores could range from 0 to 100.

### Computational model

To model behavior on the heartbeat tapping task, we first divided each task time series into intervals corresponding to the periods of time directly before and after each heartbeat. Potentially perceivable heartbeats were based on the timing of the peak of the electrocardiogram (EKG) R-wave (signaling electrical depolarization of the atrioventricular neurons of the heart) + 200 milliseconds (ms). This 200 ms interval was considered a reasonable estimate of participants’ pulse transit time (PTT) according previous estimates for the ear PTT (74). We also measured the average PTT of each participant during a separate resting-state period, defined as the distance between the peak of the EKG R-wave and the onset of the peak of the PPG waveform (signaling mechanical transmission of the systolic pressure wave to the earlobe). These average PTT values were used as covariates in some analyses (described below), and the group-level average PTT value was imputed for individuals where PPG data quality was poor. The length of each heartbeat interval (i.e., the “before-beat interval” and “after-beat interval”) depended on the heart rate. For example, if two heartbeats were 1 second apart, the “after-beat interval” would include the first 500 ms after the initial beat and the “before-beat interval” would correspond to the 2^nd^ 500 ms. The after-beat intervals were considered the time periods in which the systole (heart muscle contraction) signal was present and in which a tap should be chosen if it was felt. The before-beat intervals were treated as the time periods where the diastole (heart muscle relaxation) signal was present and in which tapping should not occur (i.e., assuming taps are chosen in response to detecting a systole; e.g., as supported by (75)). This allowed us to formulate each interval as a “trial” in which either a tap or no tap could be chosen and whether a systole or diastole signal was present (see **Figure 1**).

**Figure 1.**
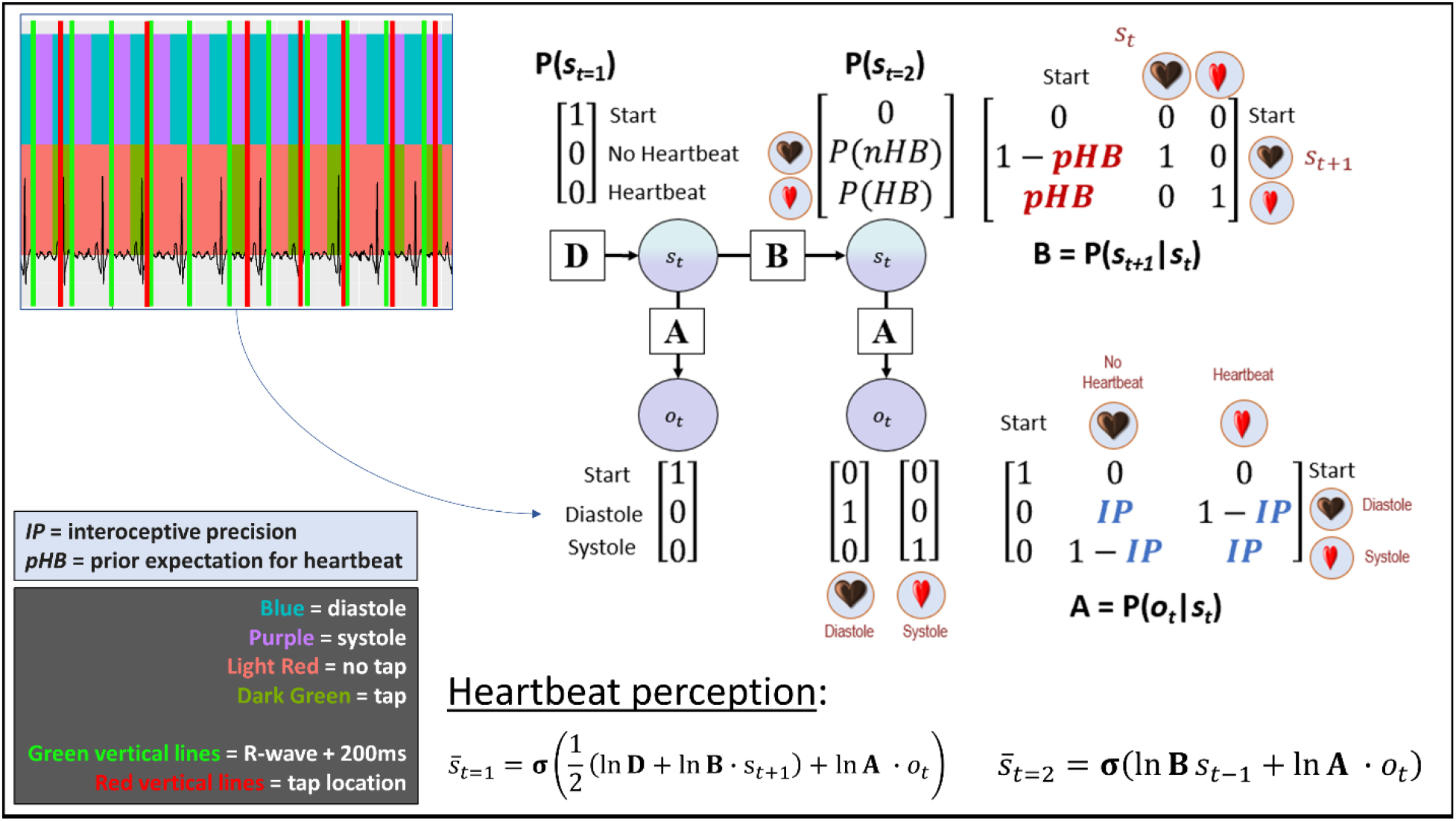
Bayesian approach used to model interoceptive awareness on the heartbeat tapping task. The generative model is here depicted graphically, such that arrows indicate dependencies between variables. Associated vectors/matrices are also shown. At each time point (t), observations (o) depend on hidden states (s), where this relationship is specified by the **A** matrix, and those states depend on previous states (as specified by the **B** matrix, or the initial states specified by the **D** vector). This model represents a simplified version of a commonly used active inference formulation of partially observable Markov decision processes (for more details regarding the structure and mathematics describing these models, see (76-78). In this model, the observations were systole/diastole, and the hidden states included beliefs about the presence or absence of a heartbeat. Selection of the tap vs. no tap actions were sampled from the posterior distribution over states 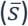 – that is, a higher posterior probability of a heartbeat state (*P*(*HB*)) corresponded to a higher probability of choosing to tap, and a higher posterior probability of the no heartbeat state (*P*(*nHB*)) corresponded to a higher probability of choosing not to tap. The model parameters we estimated corresponded to: 1) interoceptive precision (IP) – the precision of the mapping from systole/diastole to beliefs about heartbeat/no heartbeat in the **A** matrix, which can be associated with the weight assigned to sensory prediction errors; and 2) prior beliefs for the presence of a heartbeat (pHB). Because minimal precision corresponds to an IP value of .5, and both higher and lower values indicate that taps will more reliably track systoles (albeit in an anticipatory or reactive manner), our ultimate measure of precision subtracted 0.5 from raw IP values and then took their absolute value. The raw IP values were then used to assess for group differences in the tendency to tap before vs. after each systole. We also compared this model to an analogous model that included learning (see main text). On each trial, beliefs about the probability of a heartbeat (corresponding to the probability of choosing to tap) relied on Bayesian inference as implemented in the “heartbeat perception” equations shown at the bottom of the figure. Note that, by convention in the active inference models from which our model was derived, the dot product (·) applied to matrices here indicates transposed matrix multiplication. Observed diastoles and systoles were taken from ECG traces, after dividing the time series into periods before and after each systole (exemplified in the upper left portion of the figure; see text for details). Each time period in which a diastole or systole could be present was treated as a separate trial, in which the participant started in the ‘start’ state and then updated beliefs about hidden states based on observation of a diastole or systole. For this reason, the pHB parameter in the transition matrix (**B**) only specifies the probability of transitioning from the ‘start’ state to the heartbeat vs. no heartbeat states, and the heartbeat vs. no heartbeat states simply have identity mappings (i.e., a given trial cannot transition between these two states).

To model interoception and task behavior, we used a Bayesian model of perception derived from a Markov decision process (MDP) formulation of active inference that has been used in previous work; for more details about the structure and mathematics of this class of (discrete state space) models, see (77, 79, 80). See **Figure 1** for a graphical depiction of our model and the associated vectors and matrices. Observations (*o*) in the model were categorical and included systole, diastole, and a “start” observation. Hidden states (*s*) in the model, which were inferred based on observations, were also categorical and included a heartbeat state, a no heartbeat state, and a “start” state. Each trial in the model corresponded to a window in the EKG time series in which either a systole or diastole signal was present. Each trial formally had two timesteps (*t*=1 and *t*=2). At *t*=1, the participant always formally began in the “start” state and made the associated “start” observation. At *t*=2, the participant either observed a systole or a diastole and inferred whether they had transitioned from the “start” state into the heartbeat state or the no heartbeat state. In other words, they inferred a posterior distribution over states 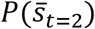 that assigned a probability to the heartbeat state and to the no heartbeat state, where this posterior distribution was informed by 1) prior beliefs about the probability of transitioning from the “start” state to the heartbeat state vs. the no heartbeat state, *P*(*s_t=_*_2_*|s_t=_*_1_), and 2) beliefs about the likelihood of observing a systole or diastole given a heartbeat or no heartbeat state, *P*(*o_t_*|*s_t_*).

A vector **D** encoded prior beliefs over initial states, *P*(*s_t=_*_1_), which specified that the participant always started the trial in the “start” state with a probability of 1 (see vector in upper left portion of the model depiction in **Figure 1**). A matrix **B** encoded the probability that each state would transition into any other state:

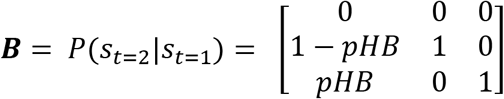

Here, columns indicate (from left to right) the “start” state, the no heartbeat state, and the heartbeat state at time *t*=1, and rows (from top to bottom) indicate the “start” state, the no heartbeat state, and the heartbeat state at time *t*=2. The probability of transitioning from the “start” state to a heartbeat state vs. a no heartbeat state was encoded by a parameter pHB, where values above 0.5 indicate prior beliefs that transitions from the “start” state to the heartbeat state are more likely (e.g., expecting a faster heart rate), and values below 0.5 indicate prior beliefs that transitions from the “start” state to the heartbeat state are less likely (e.g., expecting a slower heart rate). Note that the second and third columns simply indicate that, once entering a heartbeat or no heartbeat state, this does not subsequently change within the trial (i.e., as subsequent systole/diastole observations are modelled as subsequent trials).

A matrix **A** encoded the probability of observations given states:

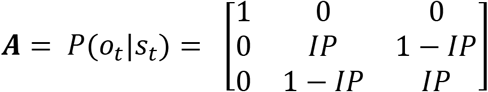

Here, columns indicate (from left to right) the “start” state, the no heartbeat state, and the heartbeat state, and rows (from top to bottom) indicate the “start” observation, the diastole observation, and the systole observation. The probability of observing a systole or diastole if a heartbeat or no heartbeat state were present was encoded by an “interoceptive precision” parameter (IP). A value of 0.5 for IP indicates minimal precision – that is, that the probability of observing a systole or diastole is 0.5 when in either the heartbeat or no heartbeat state. In contrast, a value approaching 1 indicates high precision – that is, that the probability of observing a systole is high when in a heartbeat state and low when in a no heartbeat state (and vice-versa when observing a diastole). Importantly, however, IP values approaching 0 also indicate high precision, in that observing a systole vs. diastole still provides strong evidence for a heartbeat vs. a no heartbeat state (but where the probabilistic relationships are reversed). Behaviorally, IP values approaching 0 would indicate that a participant consistently tapped the button during the task in a way that reliably anticipated each upcoming systole. It is important to note here that ‘precision’ in discrete state-space models, such as the one used here, takes on a formally different meaning than in continuous state-space models, as can be seen in the likelihood (**A**) matrix specified above. While precision refers to inverse variance in continuous state-space models, in discrete models it more simply reflects how peaked vs. flat the probabilistic mapping is between states and observations.

Belief updating in the perception model was based on the following equations at times *t* = 1 and *t* = 2, respectively:

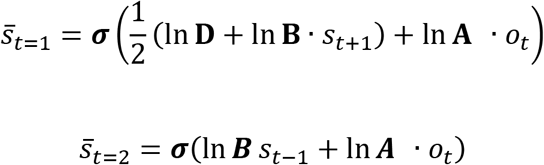

Because time *t* = 1 in each trial always included the “start” observation with a fully precise prior belief in **D** for being in the “start” state, the posterior belief 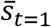 always corresponded to a fully precise belief of being in the “start” state. The equation for time *t* = 2 corresponds to Bayesian inference, in which prior beliefs (ln**B***s_t_*_−1_) are integrated with the likelihood distribution and the systole or diastole observation at the second timepoint (ln ***A*** *· o_t_*), and then converted into a proper probability distribution via a softmax (normalized exponential) function ***σ***(·) *–* leading to a posterior distribution over heartbeat states 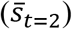. Note that, by convention in the active inference literature, the dot notation (·) here indicates a matrix transpose, meaning that **A** *· o_t_* = **A*^T^****o_t_*.

Our response model formally included two actions, the choice to tap or not tap. This model made the assumption that the probability of choosing to tap vs. not tap corresponded to the posterior probability assigned to the heartbeat vs. no heartbeat state at time *t*=2 in each trial:

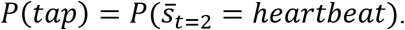

In other words, tapping behaviors were sampled from the posterior distribution over heartbeat vs. no heartbeat states, such that choices to tap became more likely as the posterior probability of a heartbeat state approached 1 and choices not to tap became more likely as the posterior probability of a heartbeat state approached 0. No further parameters were included in the response model to account for behavioral stochasticity. This is because, in the context of the present task, parameters encoding randomness in behavior cannot be distinguished from IP, as both effectively control the precision of the posterior distribution from which tapping actions are sampled in response to the systole/diastole signal. As described further below, we instead took further analysis steps at the group level to account for the potential influence of individual differences in motor stochasticity on IP and pHB estimates.

**Table 2.**
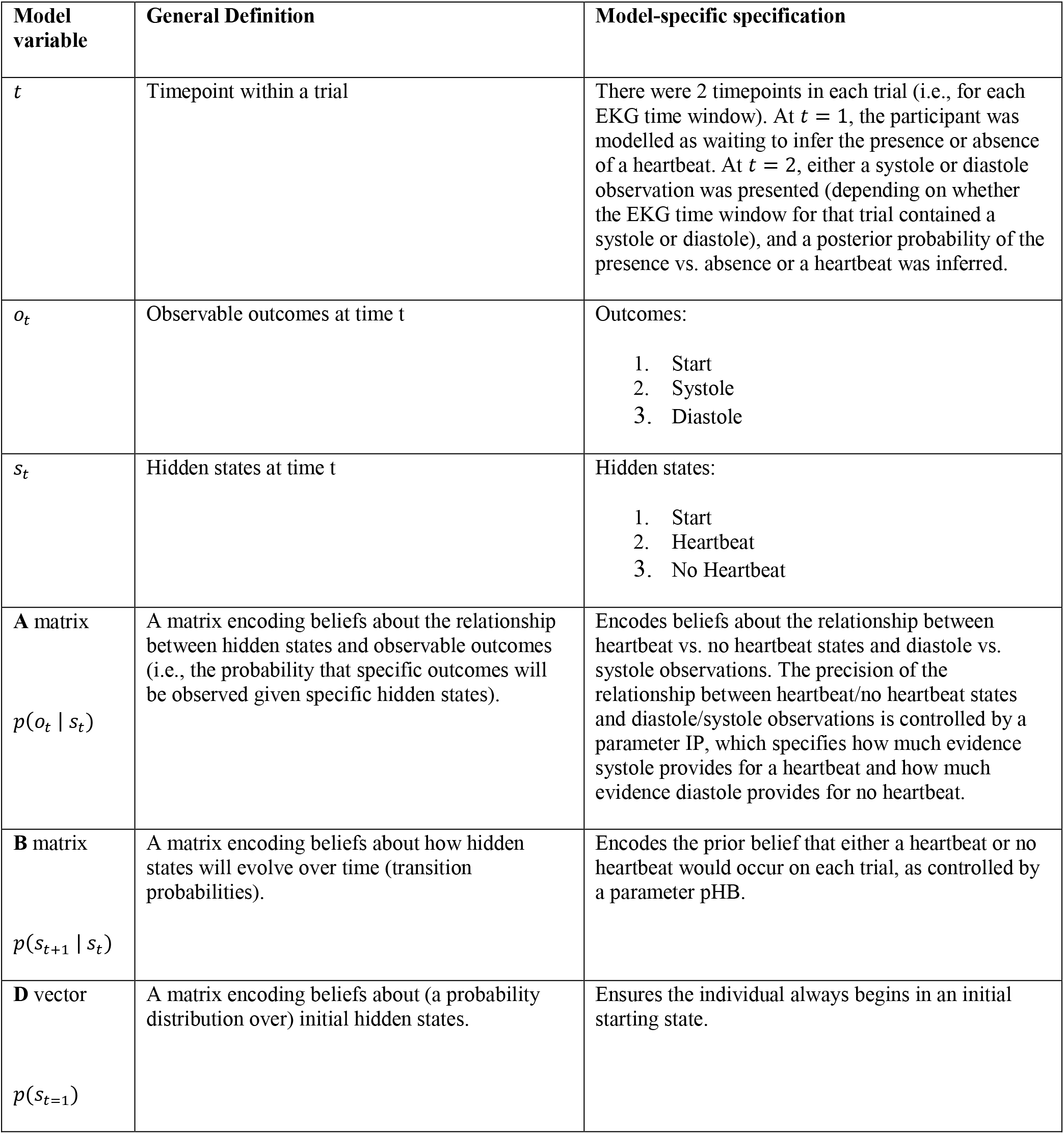
Description of computational model elements

In this task, it is unclear whether or not learning over time (e.g., from paying attention to one’s heartbeat) contributes to task performance. To assess this using Bayesian model comparison, we compared evidence for the “perception only” model described above with evidence for a model that included learning to update prior beliefs over time. Learning within this model involves updating beliefs about the probability of feeling a heartbeat vs. no heartbeat at each time point, based on how frequently one believes they have felt their heartbeat in the past. Essentially, every time a heartbeat is felt, prior beliefs favoring feeling a heartbeat go up, and every time no heartbeat is felt this (relative) belief goes back down. Formally, this corresponds to updating the concentration parameters of Dirichlet (Dir) priors associated with the **B** matrix (*b*) that specify beliefs about state transitions. At *t* = 0:

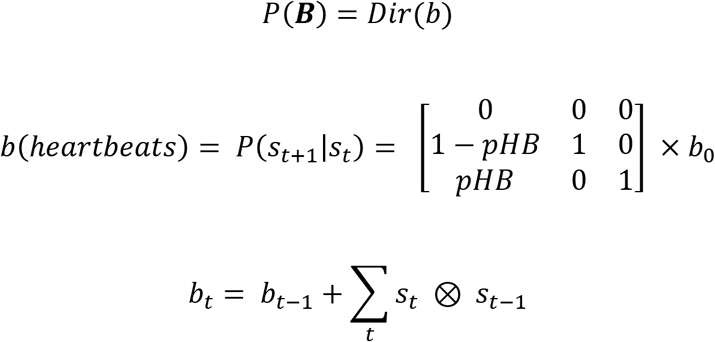

Here ⊗ indicates the cross-product, and *b*_0_ is a scalar on the prior value for concentration parameters, where its value prior to learning encodes (inverse) sensitivity to information, such that higher values will reduce the rate at which prior beliefs are updated over time with new observations. In the learning model, *b*_0_ was also estimated for each individual to capture the possibility of different learning rates for updating prior beliefs (this could also be thought of as differences in a kind of interoceptive “belief rigidity”).

Thus, the final parameters estimated for each participant included the IP, pHB, and *b*_0_ parameters. Our approach to parameter estimation used Bayesian inference at two levels (81). First, each participant’s responses were modeled using the Bayesian model of perception described above. We then used a commonly used Bayesian optimization algorithm (called Variational Bayes) to estimate each participant’s parameter values that maximized the likelihood of their responses (under the assumption that a higher/lower probability assigned to feeling a heartbeat corresponded to a higher/lower probability of choosing to tap), as described in (25). We optimized these parameters for each model using this likelihood and variational Laplace (82), implemented within the spm_nlsi_Newton.m parameter estimation routine available within the freely available SPM12 software package (Wellcome Trust Centre for Neuroimaging, London, UK, http://www.fil.ion.ucl.ac.uk/spm). This estimation approach has the advantage of preventing overfitting, due to the greater cost it assigns to moving parameters farther from their prior values. Estimating parameters required setting prior means and prior variances for each parameter. The prior variance was set to a high precision value of 1/2 for each parameter (i.e., deterring overfitting), and the prior means were set as follows: IP = .5, pHB = .5, and *b*_0_ = 1. Our decision for selecting these priors was motivated in part by initial simulations confirming that parameter values were recoverable under these prior values. The IP and pHB prior values were further chosen to minimize estimate bias, as pHB = .5 assumes flat prior beliefs, and IP = .5 does not bias estimates in favor of values assuming anticipatory vs. reactive strategies. The *b*_0_ prior of 1 is equivalent to this parameter having no effect on the model. After fitting parameters for each model, we then performed Bayesian model comparison (based on (83, 84)) to determine the best model. We then used classical inference to test for the effects of group differences in parameter estimates (i.e., posterior mean estimates) for the best model, using a standard summary statistic approach.

Before using these parameters in further analyses, however, the “raw” IP parameter values (*IP_raw_*) were transformed to correctly capture 2 distinct constructs of interest. First, because *IP_raw_* values both above and below .5 indicate higher precision (i.e., values below approaching 0 indicate reliable anticipatory tapping, whereas values approaching 1 indicate reliable tapping after a systole), our ultimate measure of precision was recalculated by centering *IP_raw_* on 0 and taking its absolute value as follows:

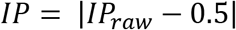

This means that IP has a minimum value of 0 and a maximum value of 0.5. The *IP_raw_* values were then instead used to assess individual differences in the tendency to tap in an anticipatory or reactive (*AvR*) fashion:

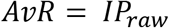

Higher AvR values (> 0.5) thus indicated a stronger tendency to reactively tap in response to a heartbeat as opposed to tapping in an anticipatory fashion (< 0.5).

### Physiological measurements

Electrocardiography was used to assess the objective timing of participants’ heartbeats throughout the task. A BIOPAC MP150 was used to collect a three lead EKG signal and the pulse oximeter signal, using a pulse plethysmography (PPG) device attached to the ear lobe. Response times were collected using a task implemented in PsychoPy, with data collection synchronized via a parallel port interface.

EKG and response data were scored using in-house developed MATLAB code. As described above in relation to modelling, each participant’s pulse transit time (PTT) was estimated as the median delay between R wave and the corresponding inflection in the PPG signal. **Supplementary materials** display an example participant’s EKG and PPG trace to illustrate how this was calculated (also supporting the 200ms delay assumption used in modelling and in individuals without usable PPG).

### Quality Control and Final Sample Sizes

Prior to performing our analyses, several participants were removed due to quality control checks: 19 individuals were removed due to “cheating” (i.e., video revealed they were taking their pulse while performing the task); 3 individuals didn’t complete the task; and 13 individuals had poor EKG across all trials that didn’t allow reliable identification of heartbeat timing. An additional 31 individuals were removed due to being outliers when performing the tone task (using Iterative Grubb’s with p < 0.01) – assumed to reflect inappropriate engagement during the task (e.g., being inattentive, tapping rapidly without listening to the tones, etc.). This resulted in 434 participants, including 52 HCs, 15 ANX, 69 DEP, 153 co-morbid ANX/DEP, 131 SUDs, and 14 eating disorders. In the three heartbeat tapping conditions, a few other participants were removed due to poor EKG, leading to the following participant numbers in each condition: 433 (guessing), 431 (no-guessing), and 427 (breath-hold). **Table 3** reports the final number of participants included for model-based analyses in each condition as well as descriptive statistics for all task-related measures.

**Table 3.** Summary statistics for all task-related variables.

Panel A.
| **<u>Guessing Condition</u>** | **Healthy comparison s (N = 52)** | **Anxiety (N = 15)** | **Depression (N = 69)** | **Depression + Anxiety (N = 153)** | **Eating Disorder (N = 14)** | **Substance use disorder (N = 130)** | **p-value\*** |
| --- | --- | --- | --- | --- | --- | --- | --- |
| **Number of taps** | 67.92 (29.58) | 58.40 (28.40) | 67.65 (31.97) | 62.02 (32.58) | 65.86 (27.86) | 65.50 (30.82) | 0.701 |
| **Number of heartbeats** | 66.67 (9.48) | 67.13 (9.55) | 70.23 (10.90) | 70.34 (10.86) | 67.21 (8.71) | 71.23 (10.47) | 0.1 |
| **Self-reported difficulty** | 56.87 (22.70) | 53.20 (26.69) | 57.55 (28.12) | 53.18 (25.84) | 71.57 (17.78) | 48.24 (24.65) | 0.009 |
| **Self-reported confidence** | 32.67 (19.27) | 41.27 (21.54) | 39.52 (22.67) | 39.29 (21.20) | 25.14 (21.36) | 42.20 (22.39) | 0.022 |
| **Self-reported intensity** | 20.71 (16.72) | 26.73 (21.51) | 26.26 (21.52) | 28.27 (23.25) | 15.36 (18.64) | 35.94 (24.65) | <0.001 |
| **Counting accuracy** | 0.69 (0.33) | 0.67 (0.30) | 0.65 (0.29) | 0.63 (0.30) | 0.68 (0.30) | 0.67 (0.29) | 0.755 |

\*p-values correspond to the results of ANOVAs comparing the groups (i.e., not including all task conditions within the analysis as reported in the main text).

Panel B.
| **<u>No-Guessing Condition</u>** | **Healthy comparison s (N = 52)** | **Anxiety (N = 15)** | **Depression (N = 68)** | **Depression + Anxiety (N = 153)** | **Eating Disorder (N = 14)** | **Substance use disorder (N = 129)** | **p-value** |
| --- | --- | --- | --- | --- | --- | --- | --- |
| **Number of taps** | 18.73 (22.69) | 22.13 (24.04) | 23.43 (24.22) | 24.10 (24.26) | 18.29 (21.74) | 31.47 (25.46) | 0.015 |
| **Number of heartbeats** | 65.88 (10.22) | 66.73 (11.07) | 69.97 (10.45) | 69.78 (11.10) | 66.14 (8.43) | 70.69 (10.14) | 0.066 |
| **Self-reported difficulty** | 67.83 (27.94) | 59.73 (35.14) | 68.00 (32.72) | 62.88 (31.35) | 69.86 (26.68) | 55.14 (30.75) | 0.039 |
| **Self-reported confidence** | 39.94 (30.32) | 36.80 (35.54) | 41.15 (31.36) | 41.41 (29.49) | 31.93 (32.16) | 42.29 (28.27) | 0.858 |
| **Self-reported intensity** | 18.15 (19.63) | 16.80 (22.11) | 25.82 (26.38) | 25.31 (25.67) | 13.50 (13.51) | 33.64 (27.53) | 0.001 |
| **Counting accuracy** | 0.28 (0.29) | 0.31 (0.33) | 0.32 (0.31) | 0.33 (0.31) | 0.28 (0.32) | 0.43 (0.32) | 0.028 |

Panel C.
| **Breath Hold Condition** | **Healthy comparisons (N = 52)** | **Anxiety (N = 15)** | **Depression (N = 68)** | **Depression + Anxiety (N = 151)** | **Eating Disorder (N = 14)** | **Substance use disorder (N = 127)** | **p-value** |
| --- | --- | --- | --- | --- | --- | --- | --- |
| **Number of taps** | 27.12 (21.47) | 30.60 (42.93) | 24.75 (21.07) | 26.46 (23.58) | 17.71 (16.80) | 34.17 (25.05) | 0.027 |
| **Number of heartbeats** | 69.40 (9.80) | 66.87 (12.01) | 71.26 (10.48) | 70.59 (11.20) | 66.29 (6.29) | 72.96 (10.75) | 0.061 |
| **Self-reported difficulty** | 46.44 (27.08) | 54.20 (23.24) | 56.53 (25.74) | 53.50 (28.32) | 55.64 (25.67) | 45.86 (29.08) | 0.07 |
| **Self-reported confidence** | 47.79 (26.20) | 41.27 (26.06) | 50.78 (26.49) | 52.42 (27.42) | 36.50 (33.37) | 53.04 (25.55) | 0.158 |
| **Self-reported intensity** | 38.19 (28.40) | 26.40 (23.61) | 41.85 (29.60) | 41.09 (29.99) | 32.71 (30.63) | 46.81 (28.88) | 0.074 |
| **Counting accuracy** | 0.39 (0.29) | 0.28 (0.26) | 0.34 (0.27) | 0.35 (0.29) | 0.28 (0.29) | 0.44 (0.30) | 0.03 |

Panel D.
| **Tone Condition** | **Healthy comparisons (N = 52)** | **Anxiety (N = 15)** | **Depression (N = 69)** | **Depression + Anxiety (N = 153)** | **Eating Disorder (N = 14)** | **Substance use disorder (N = 131)** | **p-value** |
| --- | --- | --- | --- | --- | --- | --- | --- |
| **Number of taps** | 77.90 (1.00) | 77.40 (1.12) | 77.71 (1.03) | 77.82 (1.17) | 77.57 (0.94) | 78.06 (0.97) | 0.072 |
| **Number of heartbeats** | 65.63 (11.10) | 67.27 (9.81) | 70.74 (9.94) | 71.37 (11.14) | 66.86 (8.16) | 71.30 (10.62) | 0.01 |
| **Self-reported difficulty** | 18.04 (15.84) | 29.20 (21.12) | 22.14 (19.52) | 24.15 (22.00) | 35.21 (22.33) | 25.77 (22.40) | 0.065 |
| **Self-reported confidence** | 77.56 (14.26) | 73.93 (17.84) | 78.84 (16.60) | 74.22 (19.59) | 59.79 (24.84) | 75.16 (18.56) | 0.017 |
| **Self-reported intensity** | 85.58 (13.76) | 75.67 (15.19) | 86.20 (15.09) | 82.86 (17.83) | 87.79 (12.46) | 81.47 (16.51) | 0.102 |
| **Counting accuracy** | 0.99 (0.01) | 0.99 (0.01) | 0.99 (0.01) | 0.99 (0.01) | 0.99 (0.01) | 0.99 (0.01) | 0.922 |

### Statistical analysis

Correlational analyses were first conducted to examine the relationships between parameters across conditions. For purposes of parameter validation, we ran further correlational analyses to examine the relationships between each parameter and task-specific measures, including the self-report ratings of difficulty, confidence, and heartbeat intensity collected after each trial.

Linear mixed effects models (LMEs; described further below) were conducted (using the lme4 package in R) to identify possible group differences (i.e., between HCs and the five patient groups) for each parameter and in how they differed between conditions (i.e., guessing, no-guessing, breath-hold), while accounting for individual differences in age, sex, BMI, median PTT, number of heartbeats (and its interaction with group and condition), and medication status (i.e., one analysis per parameter). Inclusion of the peripheral physiological variables was done to rule out the possibility that differences in parameters between groups/conditions was due to differences in peripheral physiology between groups/conditions. We also included sensory precision estimates for the tone condition in the models in order to account for any variability in tapping behavior due to motor stochasticity (e.g., differences in reaction times). This was based on the assumption that, because the sensory signal in the tone condition is highly precise, any variability in precision would be better explained by random influences on behavior as opposed to perception. Because the first half of the T1000 sample was pre-specified as an exploratory sample, we report relationships at *p* < .05 as a potential basis for a priori hypotheses verification in the second (confirmatory) half of the T1000 dataset in subsequent work. Although our analyses are exploratory, we note that a Bonferroni corrected threshold for multiple comparisons with three parameters is *p* < .017 (α=0.05).

For each parameter we ran LMEs (with the covariates mentioned above) that compared the three task conditions by group. Given our transdiagnostic focus, we first divided participants into two groups comprised of healthy participants and those diagnosed with one or more psychiatric disorders (transdiagnostic group). We subsequently divided those with psychiatric diagnoses into the 5 distinct diagnostic groups to assess the potential for diagnosis-specific differences.

To test the relative evidence for models with vs. without group and condition effects, we ran a JZS Bayes factor analyses with default prior scales in R (85, 86) comparing null models (with only an effect of subject) to the space of models including all combinations of main effects and interactions for group and condition. We also compared evidence for models with transdiagnostic vs. condition-specific effects.

## Results

### Participant characteristics

Complete information on sample size, demographics, and symptom screening measures is provided in **Table 1**. Separate ANOVAs showed significant differences between groups in age (*F(5,428)* = 2.98, *p* = .01) and body mass index (BMI; *F(5,413)* = 4.04, *p* = .001). A chi-squared analysis also showed significant differences in the proportion of males to females between groups (chi-squared = 19.43, df = 5, *p* = .002). Therefore, in our analyses of model parameters, we also confirm our results after controlling for these other factors.

### Interoceptive Perturbation Validation

As expected, across all participants self-reported heartbeat intensity, confidence in task performance, and task difficulty differed significantly between the three heartbeat tapping conditions in separate LMEs (intensity: *F(2858)* = 79.09, *p* < .001; confidence: *F(2,858) = 36.23, p* < .001; difficulty: *F(2,858)* = 31.8*, p* < .001), reflecting a greater perceived intensity of heartbeat sensations and greater confidence during the breath-hold perturbation than in the other 2 conditions, as well as lower difficulty in the breath-hold condition than the no-guessing condition (and greater difficulty in the no-guessing than guessing condition; *p* < .001 for all posthoc comparisons). An LME analysis of heart rate revealed a significant difference in the number of heartbeats between conditions (*F(2,856)* = 15.08*, p* < .001), reflecting a faster heart rate in the breath-hold condition than in the no-guessing condition (*p* < .001) and guessing condition (*p* = .001)

### Bayesian Model Comparison

When comparing models (based on (83, 84)), there was more evidence for the “perception only” model than for the model that included learning prior beliefs for the no-guessing and breath-hold conditions (protected exceedance probability = 1), whereas there was not clear evidence favoring a single model for the guessing condition (protected exceedance probabilities = .46 vs. .54, slightly favoring the learning model). The learning model was favored in the tone condition (protected exceedance probability = 1). No group differences were observed when comparing model fits between groups.

For consistency/comparability, we use the “perception only” model parameters to compare conditions in our analyses below, as this model best explained heartbeat tapping behavior overall. The accuracy of this model – defined as the percentage of choices to tap/not tap that matched the highest probability action in the model (e.g., a tap occurring when the highest probability percept in the model was a heartbeat) – was 74% across all conditions; by condition, model accuracy was: guessing condition = 67% (SD = 12%); tone condition = 67% (SD = 12%); no-guessing condition = 85% (SD = 14%); breath-hold condition = 84% (SD = 13%).

### Relationship between parameters

Parameter values for each group and condition are listed in **Table 4**. Across conditions, all parameters were normally distributed (skew < 2). As shown in **Figure 2**, correlations between IP across task conditions were generally low. Correlations between pHB estimates across conditions were moderate, most notably between the no-guessing and breath-hold conditions (which also included the no-guessing instruction). The tendency to tap in an anticipatory vs. reactionary manner (AvR) showed no relationships across conditions. Correlations between IP and pHB (or pTone) within each condition were also low (0 ≤ r ≤ .27), as were correlations between these parameters and AvR (-.28 < r < .01; see **supplementary materials**).

**Table 4.** Mean (and standard deviation) for model parameters by group and condition.

| Individual difference variable | Healthy comparisons | Anxiety | Depression | Depression + Anxiety | Eating disorder | Substance use disorder | p-value* |
| --- | --- | --- | --- | --- | --- | --- | --- |
| **Sensory precision** |  |  |  |  |  |  |  |
| Guessing | 0.04 (0.05) | 0.04 (0.02) | 0.04 (0.03) | 0.04 (0.04) | 0.05 (0.05) | 0.04 (0.04) | 0.815 |
| No-Guessing | 0.04 (0.05) | 0.04 (0.04) | 0.03 (0.04) | 0.03 (0.03) | 0.06 (0.06) | 0.04 (0.05) | 0.09 |
| Breath-hold | 0.07 (0.07) | 0.03 (0.03) | 0.04 (0.04) | 0.04 (0.04) | 0.03 (0.03) | 0.04 (0.04) | <0.001 |
| Tone | 0.18 (0.12) | 0.12 (0.11) | 0.18 (0.12) | 0.14 (0.11) | 0.21 (0.10) | 0.16 (0.11) | 0.052 |
| **Prior beliefs** |  |  |  |  |  |  |  |
| Guessing | 0.44 (0.16) | 0.37 (0.16) | 0.41 (0.17) | 0.38 (0.18) | 0.43 (0.18) | 0.40 (0.16) | 0.331 |
| No-Guessing | 0.15 (0.13) | 0.15 (0.12) | 0.17 (0.13) | 0.17 (0.14) | 0.14 (0.12) | 0.21 (0.14) | 0.05 |
| Breath-hold | 0.18 (0.11) | 0.19 (0.19) | 0.17 (0.11) | 0.18 (0.13) | 0.14 (0.11) | 0.22 (0.13) | 0.056 |
| Tone | 0.50 (0.01) | 0.50 (0.01) | 0.50 (0.01) | 0.50 (0.01) | 0.50 (0.01) | 0.50 (0.01) | 0.139 |
| **<u>Anticipate vs. React</u>** |  |  |  |  |  |  |  |
| Guessing | 0.49 (0.06) | 0.50 (0.04) | 0.49 (0.05) | 0.49 (0.06) | 0.54 (0.07) | 0.50 (0.06) | 0.042 |
| No-Guessing | 0.50 (0.07) | 0.52 (0.06) | 0.49 (0.05) | 0.50 (0.05) | 0.50 (0.08) | 0.50 (0.06) | 0.683 |
| Breath-hold | 0.49 (0.10) | 0.49 (0.04) | 0.51 (0.06) | 0.50 (0.06) | 0.48 (0.04) | 0.50 (0.05) | 0.529 |
| Tone | 0.50 (0.22) | 0.45 (0.16) | 0.52 (0.21) | 0.49 (0.18) | 0.37 (0.20) | 0.46 (0.19) | 0.088 |

\*p-values correspond to the results of exploratory ANOVAs comparing the groups (i.e., not including all task conditions within the analysis as reported in the main text).

**Figure 2.**
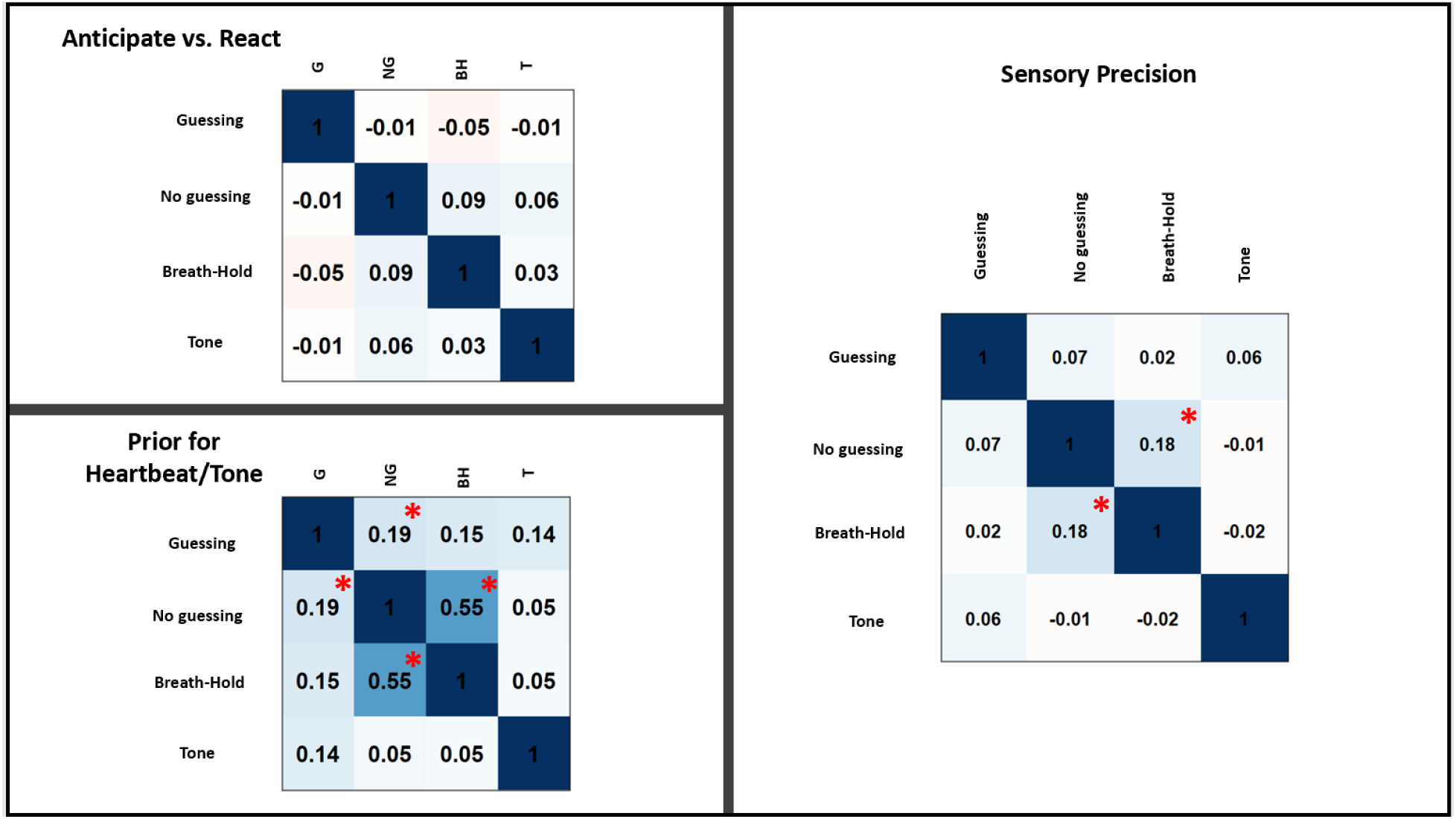
Pearson correlations between model parameters across task conditions across all participants. For reference, correlations at p < .001 (uncorrected) are marked with red asterisks.

### Parameter face validity

**Figure 3** shows the correlations, including some significant relationships (*p* ≤ .001, uncorrected), across all participants between model parameters in each condition and several task-relevant variables. IP showed positive relationships with self-reported heartbeat intensity ratings in both the no-guessing and breath hold conditions. Additionally, pHB was lower in those self-reporting greater difficulty in the no-guessing condition, and higher in those self-reporting higher confidence and higher heartbeat intensity in both the no-guessing and breath hold conditions. Across heartbeat tapping conditions, model parameters were also weakly (IP) to strongly (pHB) related to the traditional counting accuracy measure (39).

**Figure 3.**
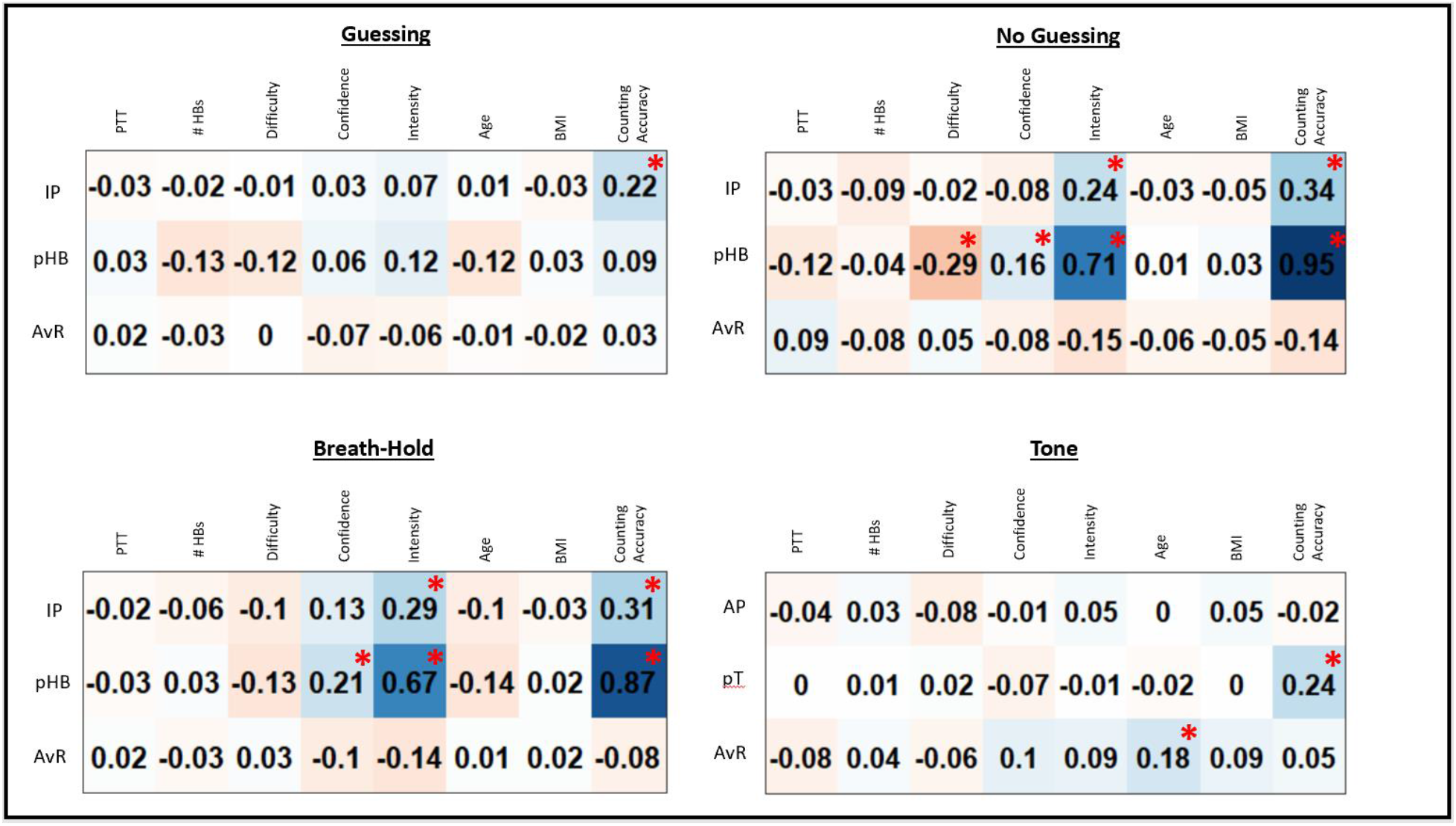
Exploratory Pearson correlations between model parameters and self-report and other taskrelevant variables for each task condition across all participants. IP = interoceptive precision parameter, pHB = prior belief for heartbeat parameter, pT = prior belief for tone parameter, AvR = anticipate vs. react strategy parameter, PTT = median pulse transit time, #HBs = number of heartbeats during the task condition, BMI = body mass index. For reference, correlations at p < .001 (uncorrected) are marked with red asterisks.

In the breath-hold condition, t-tests revealed significantly greater IP in males than females (*t*(251) = 2.30, *p* = .02).

### Group by condition interactions in model parameters

In this section, we present LME analyses assessing the main effects of task condition for each parameter, and then subsequent LMEs that exclude the tone condition, in which we assess the main effects of group and task condition, and their interaction, while accounting for several covariates that account for potential confounding effects of other demographic, behavioral, and peripheral physiological variables (see methods). In LMEs including BMI as a covariate, 15 participants were removed due to lack of available data (2 HCs, 1 ANX, 3 DEP, 1 DEP+ANX, 7 SUDs, and 1 ED).

Due to our transdiagnostic focus, we examined the transdiagnostic vs. condition-specific nature of these effects both when dividing participants into two groups (healthy comparisons vs. patients transdiagnostically) and six groups (healthy comparisons and each of the five patient groups separately). We then perform Bayes factor analyses to compare evidence for the space of possible models with vs. without group or task effects, including models with transdiagnostic effects, and models with condition-specific effects (see methods). As this is a pre-defined exploratory sample, we report post-hoc comparisons as significant with an uncorrected threshold of *p* < .05.

### Interoceptive Precision

An initial LME revealed that sensory precision (IP or auditory precision in the tone condition) was significantly different between conditions (*F(3,1292)* = 339.4, *p* < .001; see **Figure 4**), reflecting greater values in the tone condition than in the three heartbeat tapping conditions (post-hoc comparisons indicated *p* < .001 in all cases). Subsequently focusing only on IP (excluding the tone condition), and grouping participants transdiagnostically (i.e., based on the presence vs. absence of a diagnosis), a further LME revealed a significant group by condition interaction (*F(2,839)* = 10.30*,p* < .001), but no main effects of group (*F(1,607)* = 0..54, *p* = .46) or condition (*F(2,885)* = 1.35, *p* = .26). Post-hoc comparisons indicated that the interaction was driven by: 1) the fact that IP significantly increased in HCs in the breath-hold condition when compared to the two resting conditions (*p* < .001 for each), 2) that IP in HCs during breath-hold was significantly greater than IP in all three conditions in the transdiagnostic patient sample (*p* < .001 in all cases), and 3) that there were no significant differences between conditions in the patient sample. This LME also revealed no significant effect of age, sex, precision within the tone condition, BMI, median PTT, medication status, number of heartbeats, and the interaction between number of heartbeats and both group and condition (*Fs* between .02 and 1.7, *ps* between .19 and .88).

**Figure 4.**
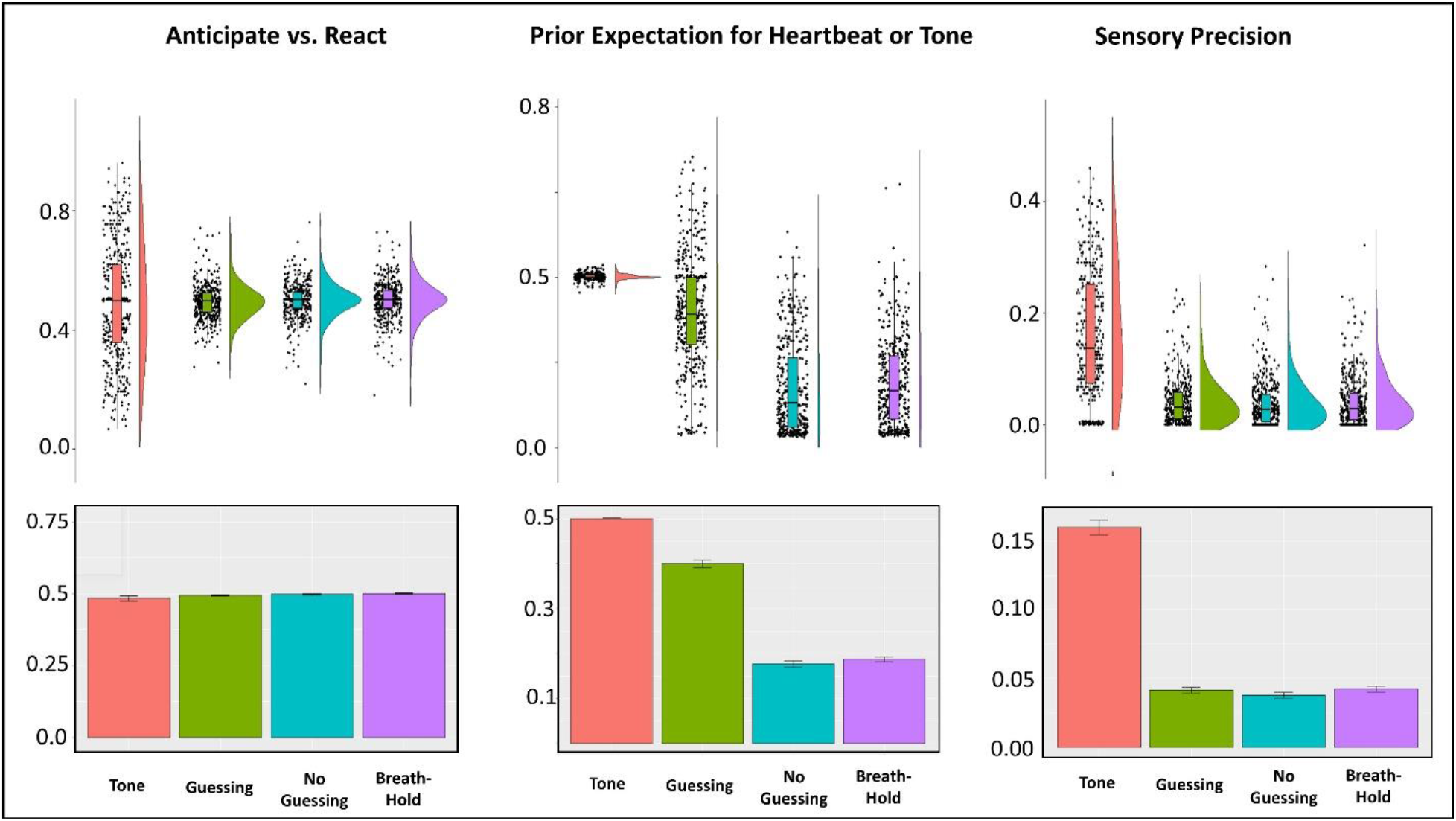
Bottom: Bar plots illustrating means and standard errors for model parameters by task condition across all participants. Prior beliefs for heartbeats were lower in the no-guessing and breath-hold conditions. Sensory precision (i.e., interoceptive precision for the heartbeat or auditory precision for the tone condition) was much greater in the tone condition. This was expected given the unambiguous nature of this signal relative to the heartbeat signal. There were no significant differences in sensory precision between the heartbeat conditions. The Anticipate vs. React values revealed no mean differences between conditions. Top: For more complete data characterization, we also show raincloud plots depicting the same results in terms of individual datapoints, boxplots (median and upper/lower quartiles), and distributions. These illustrate that, for the Anticipate vs. React parameter, nearly equally sized clusters of participants appeared to adopt more anticipatory (<.5) vs. reactive (>.5) strategies in the tone condition, and that prior beliefs remained unbiased (.5; with little variance) in the tone condition relative to the heartbeat tapping conditions.

A further LME including each diagnostic group separately confirmed the same group by condition interaction (*F(10,821)* = 3.21*,p* < .001). Here, post-hoc comparisons showed that IP in the breath-hold condition was significantly greater in HCs when compared to each clinical group (ANX, *p* = .002; DEP, *p* < .001; DEP+ANX, *pp* < .001, ED, *p* = .007, SUDs, *p* < .001). No other clinical group showed a significant difference between conditions, with the exception of DEP+ANX, which showed lower IP in the no-guessing condition than in the guessing condition (*p* = .03). DEP also showed lower IP than ED in the no-guessing condition (*p* = .03).

In the Bayesian analyses, the transdiagnostic model (HCs vs. all patients) that included main effects of group, condition, and their interaction, was most strongly favored over the null model (Bayes factor = 142.5), while the disorder-specific model was not favored over the null model (Bayes factor = .18). The transdiagnostic model was also strongly favored over the diagnosis-specific model (Bayes factor = 796.7). See **Figure 5** for plots of IP values by group and condition.

**Figure 5.**
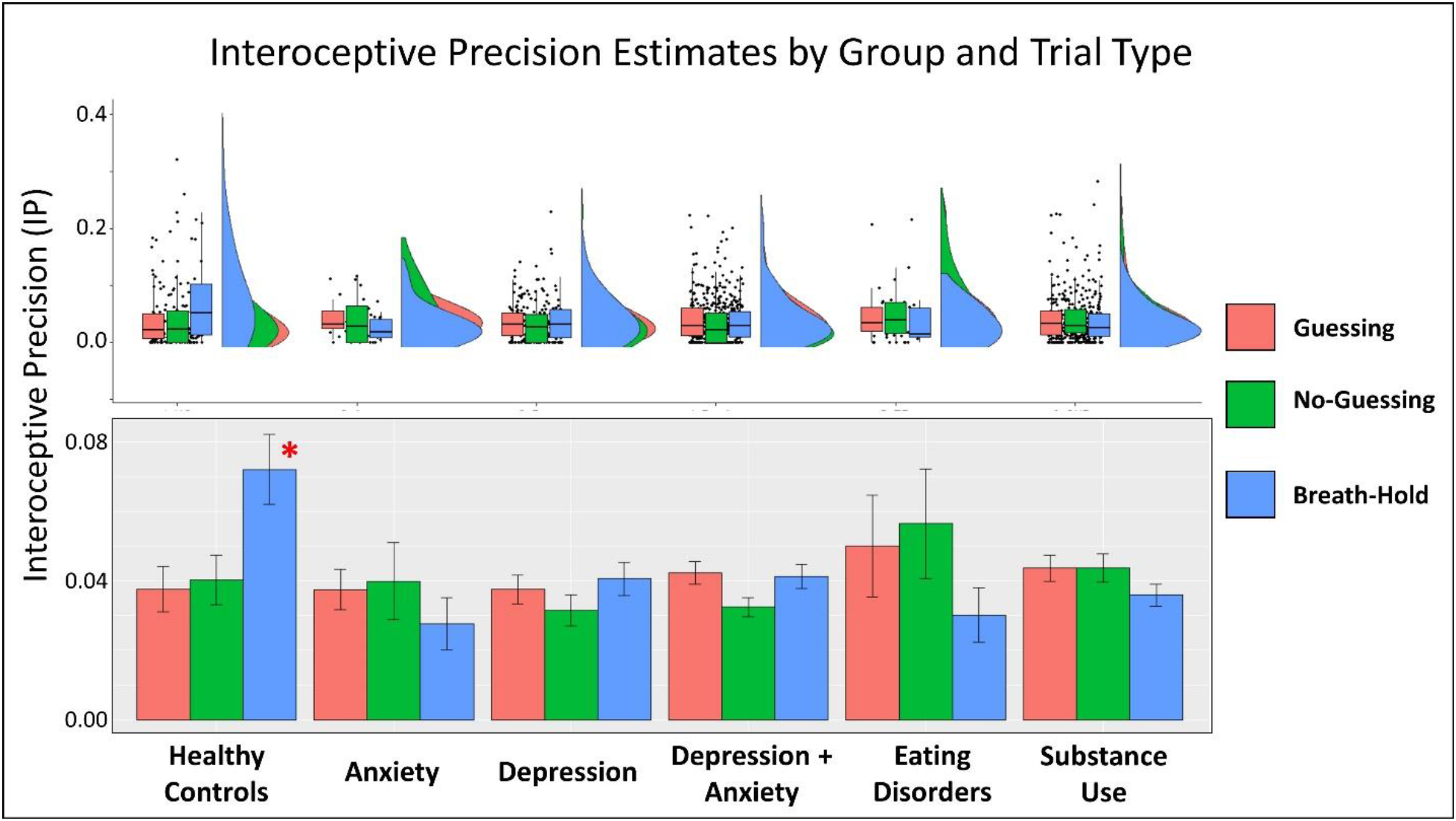
Bottom: Mean and standard error for interoceptive precision estimates by condition and clinical group. Interoceptive precision (IP) was significantly greater in healthy comparisons than all other groups in the breath-hold condition, and healthy comparisons showed a significant increase in IP from the guessing and no guessing condition to the breath-hold condition that was absent in the other groups. Top: For more complete data characterization, we also show raincloud plots depicting the same results in terms of individual datapoints, boxplots (median and upper/lower quartiles), and probability densities.

### Prior expectations

An LME revealed that prior expectations (pHB or priors in the tone condition) were significantly different between conditions (*F(3,1291)* = 861.5, *p* < .001; see **Figure 4**), reflecting greater values in the tone condition than in the three heartbeat tapping conditions, and greater values in the guessing condition than in the other two heartbeat tapping conditions (post-hoc comparisons indicated *p* < .001 in all cases). Subsequently focusing only on pHB (excluding the tone condition), and grouping participants transdiagnostically, a further LME revealed a main effect of condition (*F(2,863)* = 27.09, *p* < .001), and a group by condition interaction (*F(2,842)* = 4.38*, p* = .01), but no main effect of group (*F(1,722)* = .03*, p* = .86). Posthoc comparisons again indicated greater pHB in the guessing condition than in the other two conditions (*p* < .001). However, post-hoc comparisons did not show any significant differences between groups by condition to account for the interaction. This LME also revealed a negative association between pHB and age (*F(1,408)* = 8.84, *p* = .003) and a heart rate by condition interaction (*F(2,865)* = 3.56, *p* = .03), but no significant effect of sex, tone precision, BMI, median PTT, medication status, number of heartbeats, or the interaction between number of heartbeats and group (*Fs* between .02 and 3.49*,ps* between .06 and .90). The heart rate by condition interaction reflected a stronger negative correlation between heart rate and pHB in the guessing condition (*r* = −.13) than in the no-guessing and breath-hold conditions (*r* = −.04 and .03, respectively).

A further LME with diagnosis-specific groups confirmed the same group by condition interaction (*F(10,821)* = 2.21, *p* = .02). Here, post-hoc comparisons suggested this interaction was driven primarily by the SUD group, who showed higher pHB values in the breath-hold condition than the DEP, DEP+ANX, and ED groups (*p* = .048, .03, and .04, respectively), and also showed higher pHB values in the no-guessing condition than the DEP+ANX, ED, and HC groups (*p* = .03, .046, and .01, respectively). HCs also showed higher pHB values in the guessing condition than the DEP+ANX group (*p* = .03).

In the Bayesian analyses, the most strongly favored model over the null model only included a main effect of condition (Bayes factor = 1.07E+135). This model was also favored over 1) a model including both main effects and the transdiagnostic group by condition interaction (Bayes factor = 4.8), and 2) an analogous model with the diagnosis-specific group by condition interaction (Bayes factor = 63.21). See **Supplementary materials** for plots of pHB values by group and condition.

### Anticipating vs. reacting

An initial LME showed no significant differences in AvR between task conditions (*F(3,1292)* = 2.01, *p* = .11; see **Figure 4**). Detailed results of further LMEs and Bayes factor analyses for AvR are presented in **Supplementary materials**. No clear evidence was found for differences between HCs and the clinical groups by condition, although some potential relationships with heart rate and medication status were noted. Bayesian analyses strongly favored the null model over all other models. See **Supplementary materials** for plots of AvR values by group and condition.

### Comparison to existing measures

To examine the ability of traditional measures to capture similar group differences, we ran analogous LMEs using the traditional heartbeat counting task formula for interoceptive accuracy (39). When using the transdiagnostic grouping, analyses did not find a main effect of group (*F*(1,688) = .01, *p* = .92) or condition (*F*(2,868) = 2.4, *p* = .09), but did detect an interaction between group and condition (*F*(2,842) = 3.37, *p* = .03). There was negative association with age (*F*(1,408) = 4.59, *p* = .03) and PTT (*F*(1,412) = 4.83, *p* = .03), and no effect of any other covariate (*F*s between .0003 and 1.09, *p*s between .30 and .99). Post-hoc comparisons suggested the group by condition interaction was driven by greater counting accuracy in the guessing condition than the other two conditions in both groups (p < .001 in all cases), and greater counting accuracy in the breath-hold condition than the no-guessing condition in HCs but not in the transdiagnostic group (*p* = .045 and *p* = .55, respectively). When using the diagnosis-specific grouping, there was no main effect of group (*F*(5,620) = .23, *p* = .95) or group by condition interaction (*F*(10,821) = 1.46, *p* = .15). A Bayes factor analysis showed that the most highly favored model (relative to a null model) only included an effect of condition (Bayes factor = 1.7E+62), reflecting higher counting accuracy in the guessing condition than in the other two heartbeat tapping conditions (*p* < .001; also see **Table 3**). It was also strongly favored relative to the second-best model including an effect of group and condition (Bayes factor = 7.4).

To assess potential group differences in the effect of task condition on self-reported experience and physiology, we also carried out analogous LMEs assessing confidence, intensity, and difficulty, as well as heart rate. These results are reported in the **supplementary materials**. No group by condition interactions in these variables were observed mirroring our IP results.

### Associations with symptom severity measures and interoceptive awareness scales

Given the heterogeneity in our clinical sample, we ran subsequent exploratory correlational analyses with continuous scores on the clinical measures gathered, excluding HCs, to assess whether model parameters might provide additional information about symptom severity. We note a few weak relationships at uncorrected levels in **supplementary materials**, but none survive correction for multiple comparisons.

The T1000 dataset also includes self-report measures commonly used in interoception research, including the Multidimensional Assessment of Interoceptive Awareness (MAIA; (87)), the Toronto Alexithymia Scale (TAS-20; (88)), and the Anxiety Sensitivity Index (ASI; (89)). For the interested reader, we also show exploratory correlation matrices between model parameters and these common measures within **supplementary materials**. While a couple of relationships were significant at uncorrected levels, the strength of these relationships was low.

## Discussion

This investigation aimed to examine whether a novel Bayesian computational model of perception could provide a principled approach to empirically characterizing interoceptive processing dysfunctions that have previously been proposed. Specifically, we used behavior during a heartbeat perception task in conjunction with this model to estimate quantitative differences in the prior beliefs and sensory precision estimates that individuals implicitly apply to afferent interoceptive (cardiac) signals – in both healthy individuals and a transdiagnostic sample of individuals with depression, anxiety, substance use, and/or eating disorder symptoms. We observed several relationships in the expected directions between model parameters and other task-related variables that supported the construct validity of our model parameters.

Of greatest interest, we found evidence, using both frequentist and Bayesian analyses, that an interoceptive (breath-hold) perturbation increased the precision estimates assigned to cardiac signals in healthy individuals, but that this perturbation had no effect on interoceptive precision in individuals with a range of psychiatric disorders, including depression, anxiety, substance use, and/or eating disorders (**Figure 5**). Notably, Bayesian analyses found evidence against differences in interoceptive precision between the different clinical groups, suggesting that low precision during perturbation represents a transdiagnostic feature across these psychiatric conditions. Bayesian analyses also found evidence against differences in prior expectations between healthy participants and psychiatric patients, suggesting that differences in sensory precision, and not prior beliefs, best account for interoceptive differences in the clinical groups (although frequentist analyses suggested that substance users may show less attenuated prior expectations than some other groups in the no-guessing and breath-hold conditions). Model comparison further suggested that individuals did not update prior beliefs over time during the task, which is perhaps unsurprising given the low cardiac awareness commonly seen in previous studies (6). That is, individuals may not have had sufficient signal to learn from (note that, in contrast, model comparison supported the presence of learning in our auditory control condition with a clear signal). We expand on these points below.

### Parameter validity/sensitivity

IP showed positive relationships with self-reported heartbeat intensity ratings (no-guessing and breath-hold conditions) – as would be expected in the context of more precise cardiac signals. In the no-guessing and breath-hold conditions, pHB was lower than in the guessing condition. In other words, participants appear to have successfully adjusted their prior beliefs to comply with the no-guessing instructions. Further, consistent with the role of prior beliefs in perception, pHB was also lower in those reporting greater difficulty (no-guessing condition) and higher in those reporting greater confidence and higher heartbeat intensity (no-guessing and breath-hold conditions). Each of these results support the notion that our parameters tracked theoretically meaningful individual differences in perception/behavior and that our approach can disentangle the effects of sensory precision from those of prior beliefs and anticipatory vs. reactive strategies.

Further, while model parameters had some shared variance with traditional accuracy measures (e.g., higher counting accuracy was weakly associated with higher interoceptive precision), they mainly captured unique variance that was not tracked by standard measures. Further, no group differences analogous to those seen with model parameters were found using the traditional interoceptive accuracy measure (i.e., across all participants, counting accuracy showed an opposite pattern, being highest in the guessing condition). This highlights the unique ability of this computational method to uncover differences in perceptual decision making across different psychiatric subtypes.

It is also worth noting the strong positive correlation we observed between pHB and counting accuracy in the no-guessing and breath-hold conditions, suggesting that heartbeat counting accuracy primarily reflects prior beliefs (as previously proposed in (38)). In the present task this is explained by the fact that higher pHB values led to a greater number of taps and the fact that the average number of taps in these conditions was low (i.e., because of the no-guessing instruction). Thus, those who tapped more often (i.e., due to stronger priors, independent of precision) approached the actual number of heartbeats and therefore had higher counting accuracy scores. This highlights one specific way in which, in the context of restrictions on guessing, counting accuracy may be most closely associated with prior beliefs.

### Differences in Interoceptive Precision

Our primary results were that: 1) IP was significantly higher in the breath-hold condition in healthy comparisons than in each of the clinical groups; and 2) there was a group by condition interaction, demonstrating that the interoceptive perturbation (breath-hold) increased IP (relative to resting conditions) in the healthy participants, whereas this perturbation had no effect in any of the clinical groups. These group differences were not diagnosis-specific and were not accounted for by any other demographic (e.g., age, sex) or physiological variables (e.g., pulse transit time, changes in heart rate), suggesting it was not a result of group-specific changes in peripheral physiology between conditions. Thus, as the clinical groups had comparable IP to healthy participants during resting conditions, it appears that interoceptive processing differences in psychiatric populations may not be present across all physiological contexts. Instead, interoceptive processing differences may manifest selectively when in altered physiological states (e.g., such as the high arousal states linked with anxiety).

The finding that IP was reduced during the breath-hold across psychiatric groups included in this study may be of clinical interest. First, multiple neurocognitive (55, 90, 91) and computational (11, 12, 15, 27, 92) theories of emotion, and associated empirical findings (e.g.,(42, 93-97)), suggest that interoceptive awareness may be an important transdiagnostic factor in promoting emotional awareness. As low emotional awareness has been linked to multiple psychiatric and systemic medical conditions (reviewed in (98, 99)), reduced IP during altered interoceptive states (e.g., affective arousal) could contribute to low emotional awareness and its maladaptive consequences irrespective of diagnostic category. Second, visceral regulation might be expected to be less effective in the absence of precise feedback signals from the body, which could relate to visceral dysregulation in psychiatric conditions (e.g., see (56)). For example, to adaptively regulate visceral states, visceromotor control regions plausibly require reliable feedback signals from the body to determine whether descending visceromotor commands have successfully led to the intended adjustments in internal bodily states (10, 13, 36). Thus, the low IP observed across multiple psychiatric conditions during the breath-hold could potentially contribute to visceral dysregulation in other contexts in which interoceptive perturbations occur, such as highly arousing, negatively valenced states (e.g. panic anxiety, irritability/anger etc.).

These results build on previous bodies of work suggesting associations between psychiatric disorders and interoceptive processing deficits (38, 56, 100-103). For example, previous cardiac perception studies have shown that depressed patients exhibit reduced accuracy on a heartbeat counting task (2-4), and that performance is negatively correlated with depressive symptoms (5) as well as associated with both lower positivity and poorer decision-making (2); although, the limitations of heartbeat counting tasks should be kept in mind when interpreting such findings (47, 49, 63, 104). While the literature on interoceptive dysfunction is mixed for anxiety disorders broadly, it is well-stablished in panic disorder (reviewed in (6)). A couple recent studies have also reported evidence of differences in interoceptive processing in both SUDs (e.g., blunted brain responses (7)) and eating disorders (e.g., stronger effects of prior beliefs on perception during low arousal (8)). The computational framework within which our findings were observed is also in line with several recent proposals about the role of interoceptive inference in guiding (predictive) autonomic control and the potential breakdown of this mechanism within different psychiatric conditions (10-15, 36, 38); however, in contrast to previous emphasis on altered prior beliefs in these proposals, our results more selectively support the existence of deficits in adjusting precision estimates for afferent interoceptive signals – and do not provide strong support for the presence of altered priors.

That said, the neurobiological mechanisms promoting reduced IP in mental disorders during interoceptive perturbation remains unclear. As IP increased during the interoceptive perturbation in HCs, but not in the clinical groups, it could be that altered brain processes in individuals with certain psychiatric disorders fail to update IP estimates during states of acute bodily arousal. The neural process theory associated with active inference and related Bayesian models suggests an inability to adjust IP estimates would correspond to reductions in synaptic plasticity in response to changes in patterns of interoceptive prediction-errors (10, 24, 77, 79, 105), most plausibly within neural networks supporting interoception and visceromotor control (e.g., insula and anterior cingulate (15, 106)). This could be tested using our model/task in conjunction with neuroimaging. Alternatively, IP estimates could be accurate, and afferent interoceptive signals may in fact be conveyed with less fidelity (i.e., greater noise) to the brain in the context of psychiatric conditions. For example, such differences could be due to altered signaling in interoceptive sensory neurons, which could in principle be affected by many factors (genetic/epigenetic influences, early adversity and related socio-environmental factors, and/or effects of disease-related chronic stress, among others). Future research will need to investigate these different possibilities.

### Prior expectations and task strategy

Unlike the results for sensory precision described above, our results did not provide strong support for group differences in prior expectations or the tendency to tap in an anticipatory vs. reactive manner. While frequentist analyses suggested a few potential group differences in prior beliefs in the no-guessing and breath-hold conditions, Bayesian analyses found strong evidence favoring a model without group differences over a model with transdiagnostic or diagnosis-specific effects. This suggests that baseline expectations for heart rate do not differ in the psychiatric populations examined here, and that these priors can be adjusted in response to instructions in the same manner as in healthy participants. The lack of group differences with respect to strategy might also be seen as supporting a similar conclusion in that one might expect greater tendencies to tap in an anticipatory manner if basing behavior on prior expectations. Thus, further study of differences in the ability to adjust precision estimates may be more promising for improving our understanding of interoceptive dysfunction in psychiatric populations. However, the finding in frequentist analyses that pHB in substance users was less attenuated relative to some other groups in the no-guessing and breath-hold conditions could suggest that less sensory evidence is required to generate changes in interoceptive feelings within these individuals (consistent with the greater self-reported heartbeat intensity and lower self-reported difficulty in this group relative to the other groups suggested by our supplementary analyses; see **Table 3** and **Supplementary Materials**). This possibility is worth further investigation in future studies.

### Strengths, Limitations, and Conclusion

This study has several major strengths. First is the novel application of a computational model to behavior on an interoceptive awareness task, which allowed for model comparison (e.g., allowing us to rule out learning effects) and parameter estimates to disentangle distinct computational mechanisms (e.g., the role of prior beliefs vs. interoceptive precision). A second strength is the application of this model to interoceptive processing in individuals with psychiatric disorders, which to our knowledge, has never been reported. While accounting for other influences, this approach allowed us to test a theoretical prediction – that afferent interoceptive signals (and resulting prediction-errors) are assigned low precision estimates in those with psychiatric disorders, leading those signals to be under-weighted during interoceptive inference (although, as discussed above, other interpretations are possible). While an important first step, a further test of this hypothesis would require replication in a confirmatory sample, something we plan to investigate in the second cohort of participants in the Tulsa 1000 project (for some confirmatory work in an independent sample of healthy individuals, see (107)). Additionally, combining this approach with neuroimaging measures would help to establish the expected relationship between IP and neural activity within interoceptive cortices. Computational approaches derived from Active inference models may be especially useful in this regard, as they allow for simulation of predicted neuronal responses (79).

Some limitations of our modeling approach are also important to keep in mind. First, we were required to make specific choices about model structure. For example, while priors during estimation were fixed at plausible values (justification explained within the methods section), other values might have been chosen. Also, we chose not to include a temperature parameter in our action model. While this was possible, in simple perceptual tasks like ours that lack reward feedback, any variability in the precision of action selection would be correlated with sensory precision. Therefore, we chose a simpler model that treated behavior as a direct readout of posterior beliefs in perception. To account for potential differences in motor stochasticity, we then controlled for sensory precision in the tone condition, where, because the sensory signal is highly precise, between-subject differences in precision would most plausibly reflect between-subject differences in variability of motor responses. Here it is also worth highlighting that, if motor effects (such as reaction time) were stable within-subjects but varied between subjects, this would not influence precision estimates because the temporal relationship between systoles and taps would still be clear. However, the novelty of our approach entails that it should be replicated in future studies.

Another limitation is that, while we did compare models with vs. without learning, we did not compare our model to other existing approaches. The heartbeat tapping task was not well-suited for the several alternatives we considered. For example, it did not have a sufficient number of trials to be suited for traditional signal detection approaches, which would be most comparable to our model-based precision measure (108). Computational models based on reinforcement learning were also inappropriate as the task did not include planning or learning from reward, and instead dealt mainly with uncertainty in perception. One alternative approach we might have taken would have been to model the task with a hierarchical Gaussian filter (109), which has similar Bayesian foundations and can estimate the relative precisions of sensory input and prior beliefs (i.e., as opposed to the separate estimates of each in our model), which could be useful to examine in future work.

Our modelling approach also required making choices about how to include EKG signals as observations and relate them to behavior. Here this involved discretizing timepoints where responses were considered co-occurrent (or not) with diastole vs. systole; but other such discretizations could have been chosen. That said, given the relationships we observed between parameters and other task measures, our choices appear to have led to estimates that track meaningful individual differences in task behavior. The task measurement conditions also had certain limitations. For example, many individuals had low IP values – reflecting the low cardiac awareness commonly seen at rest in previous studies (6) – which may have limited the variability necessary to assess relationships with other variables. Finally, there were relatively lower sample sizes in the eating disorders and anxiety disorders groups. Thus, studies looking at these groups with larger sample sizes will be necessary to afford stronger confidence in our results for these groups.

In summary, this study 1) demonstrated the sensitivity of individual difference measures (parameter estimates) derived from a novel Bayesian computational model, with a focus on estimating the precision weighting of interoceptive signals across a transdiagnostic sample of individuals with psychiatric disorders, and 2) tested—and found evidence supporting—the hypothesis that individuals with psychiatric disorders fail to update the precision-weighting of afferent interoceptive signals during homeostatic perturbations. While the underlying neurophysiological mechanisms leading to this difference remain unidentified, these results point to a potential origin of visceral dysregulation (and perhaps its influence on maladaptive behavior) across multiple psychiatric conditions. This represents an important step towards a primary goal of computational psychiatry – computationally phenotyping individuals with psychiatric disorders with the tools of computational neuroscience in hopes of using this information to guide the development of precision medicine interventions.

## Data Availability

All relevant data are available within the manuscript and its Supporting Information files.

## Software Note

All model simulations were implemented using standard routines **(spm_MDP_VB_X.m)** that are available as Matlab code in the latest version of SPM academic software: http://www.fil.ion.ucl.ac.uk/spm/. The specific code used for our model can be found in supplementary materials.

## Supplementary Files

**Supplementary materials**. This file includes results of supplementary analyses as well as supplementary figures, as referred to in the main text.

**Supplementary code**. This file contains the MATLAB code used for modelling task behavior.

**Study data**. This file includes all data used in the analyses reported in the manuscript.

